# Clinical and mechanistic responses to a music-based intervention for pain in adults with irritable bowel syndrome: Protocol for a single-arm pilot study

**DOI:** 10.64898/2026.09.11.26362754

**Authors:** Weizi Wu, Eunhea You, Aolan Li, Jie Chen, Luana Colloca, AZA Allsop, Shannon Kiley, Boluwatife Faremi, Camila Jimenez Wong, John M. Toribio, Kyle J. Mahoney, Hugo Fernando Posada-Quintero, Ming-Hui Chen, Debra S Burns, Xiaomei Cong

## Abstract

Irritable bowel syndrome (IBS) is a common disorder of brain-gut interaction characterized by recurrent abdominal pain and altered bowel habits. Although music-based interventions (MBIs) have shown benefits on pain and stress, the mechanisms associated with MBI-related changes in IBS pain remain poorly understood, and the feasibility of capturing these responses using multimodal wearable monitoring in everyday settings has not been established. This protocol describes a single-arm pilot mechanistic trial examining clinical and multimodal mechanistic responses to an MBI in adults with IBS and the feasibility of extending physiological monitoring from the laboratory to home settings. Thirty-six adults aged 18-50 years with provider-confirmed IBS will be enrolled, with approximately 30 expected to complete the post-intervention laboratory visit. Participants will receive board-certified music therapist-guided laboratory sessions and complete four weeks of self-administered home practice using a standardized 20-minute MBI protocol at least five days per week. Guided by a brain-gut mechanistic framework, assessments will include patient-reported pain, stress, and IBS symptoms; quantitative sensory testing; gut microbiome profiling using 16S rRNA gene sequencing; and multimodal physiological recordings of neural, autonomic, and muscular activity. Analyses will characterize within-session and longitudinal changes in clinical and mechanistic measures and assess recruitment, retention, MBI adherence, wearable data completeness, and participant acceptability. Findings will inform the selection of candidate mechanisms and wearable monitoring approaches for evaluation in a future adequately powered randomized controlled trial.

## Introduction

Irritable bowel syndrome (IBS) is a common disorder of brain-gut interaction characterized by recurrent abdominal pain and altered bowel habits (Huang et al., 2023; Mayer et al., 2023). Recent population-based estimates indicate that approximately 6% of U.S. adults meet Rome IV criteria for IBS, which is associated with substantial individual and societal burdens, including impaired quality of life, increased healthcare utilization, reduced work productivity, and socioeconomic costs (Almario et al., 2023; Bosman et al., 2023; Nellesen et al., 2013). Although the pathophysiology of IBS remains incompletely understood, persistent abdominal pain is increasingly recognized as arising from dysregulation across interconnected components of the brain-gut axis rather than from a single peripheral gastrointestinal abnormality (Mayer et al., 2023; Salvioli et al., 2015; Zhou & Verne, 2011). Alterations in central pain processing, autonomic regulation, visceral sensitivity, psychological processes such as stress and anxiety, and the gut microbiome may collectively contribute to IBS pain and related symptoms, providing rationales for interventions that target multiple physiological and psychological processes (Goodoory et al., 2021; Grover et al., 2021; Mayer et al., 2023; Wu et al., 2025).

Current management of IBS includes pharmacological and nonpharmacological approaches yet achieving sustained relief from pain and other symptoms remains challenging. Pharmacological therapies are an important component of IBS management, but treatment response varies, and adverse effects may limit long-term use (Black & Ford, 2021; Cong, Perry, et al., 2018; Lacy et al., 2021; Lembo et al., 2022). Mind-body interventions offer a complementary nonpharmacological approach for managing IBS pain and related symptoms (Black & Ford, 2021; Orock et al., 2020). Cognitive behavioral therapy, for example, has shown benefits for IBS symptoms, psychological distress, and quality of life (Kim et al., 2022). However, broader implementation of these interventions may be constrained by the need for trained providers, treatment duration and cost, and the sustained time commitment required from patients (Orock et al., 2020). These limitations highlight the need for accessible and scalable interventions that can be readily incorporated into daily self-management.

Music-based interventions (MBIs) are one such approach because of their noninvasive nature, favorable safety profile, and accessibility (Chen et al., 2022; Lunde et al., 2019; Patiyal et al., 2021). Growing evidence suggests MBIs may improve pain, stress, anxiety, and psychological distress across a range of clinical and chronic pain populations (Cournoyer Lemaire & Perreault, 2024; de Witte et al., 2020; Lunde et al., 2019). Music-induced analgesia is thought to involve multiple interacting processes, including modulation of neural circuits involved in emotion, attention, reward, and pain processing, together with changes in physiological arousal and autonomic regulation (Arnold et al., 2024; Cannon & Patel, 2021; Chen et al., 2022; Kasdan et al., 2022; Mas-Herrero et al., 2018; Mojtabavi et al., 2020). These processes overlap with key mechanisms implicated in IBS pain, particularly altered central pain processing and autonomic regulation. Emerging preclinical evidence further suggests that music exposure may influence gut microbial composition and related biological processes, although the relevance of these findings to humans remains unclear (Niu et al., 2023; Zhang et al., 2023). Together, these findings provide rationales for examining clinical and multimodal mechanistic responses to MBIs in IBS.

However, mechanistic evidence regarding MBI-related changes in IBS pain remains limited, as previous studies have focused primarily on self-reported clinical outcomes rather than concurrent changes across central neural, autonomic, sensory, and gut-related processes (Chen et al., 2022; de Witte et al., 2020; Gantt et al., 2017). In addition, physiological responses to MBIs have typically been assessed during discrete laboratory (lab) sessions, providing limited insight into responses during repeated intervention exposure in everyday settings (Bradt et al., 2026). Advances in wearable technologies offer an opportunity to extend physiological monitoring beyond the lab and capture continuous physiological responses during home-based MBI practice, enabling characterization across both controlled and real-world settings.

To address these gaps, we designed a pilot mechanistic study of an MBI in adults with IBS. The study has two aims: (1) to characterize changes in IBS-related pain and candidate mechanistic measures, including central and autonomic physiological activity, stress responses, somatic pain sensitivity, and gut microbial profiles; and (2) to evaluate the feasibility of integrating multimodal wearable physiological monitoring into repeated home-based MBI sessions, including MBI adherence, wearable-device adherence, data completeness, and participant acceptability. Findings from this pilot study will inform the refinement of mechanistic hypotheses, measurement strategies, and study procedures for a future adequately powered randomized controlled trial.

## Methods

### Conceptual Framework

The conceptual framework (Figure 1) organizes the study around four candidate mechanistic domains: central neural processing, autonomic and stress regulation, somatic pain sensitivity, and gut microbial processes. The first three domains are hypothesized to show changes associated with MBI-related changes in IBS pain, whereas gut microbial profiles are examined as an exploratory parallel domain given the limited human evidence linking music exposure to the gut microbiome. Individual sociodemographic, clinical, and music-related characteristics are considered as potential sources of variability in these responses. The framework guides the selection and organization of study measures and the exploratory evaluation of mechanism-outcome associations.

**Figure 1.**
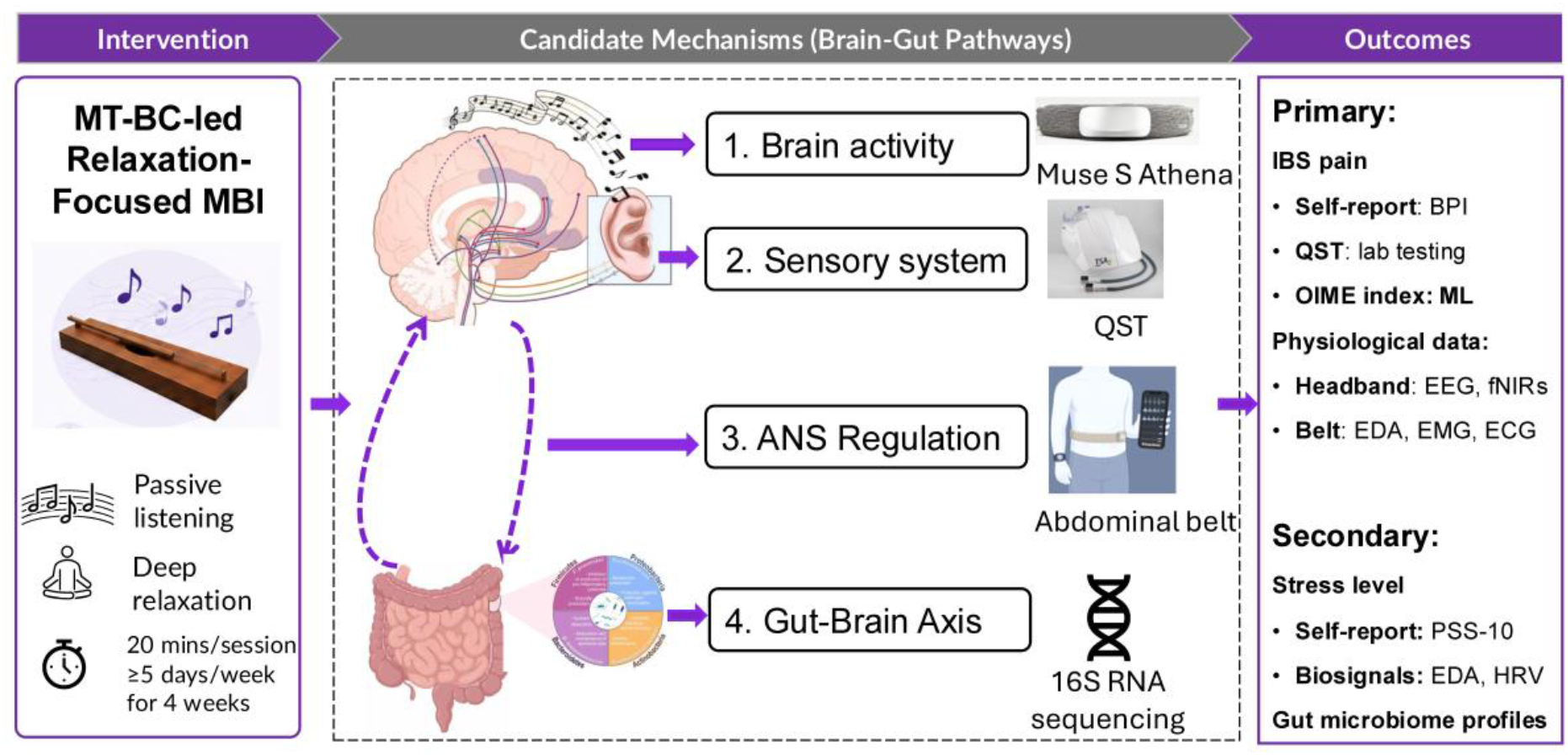
Conceptual framework of candidate mechanistic responses to the music-based intervention. The framework illustrates candidate domains hypothesized to show changes associated with MBI-related changes in IBS pain, including central neural processing, autonomic and stress regulation, and somatic pain sensitivity. Gut microbial processes are included as an exploratory parallel domain. Abbreviations: ANS, autonomic nervous system; BPI, Brief Pain Inventory; ECG, electrocardiography; EDA, electrodermal activity; EEG, electroencephalography; EMG, electromyography; fNIRS, functional near-infrared spectroscopy; HRV, heart rate variability; IBS, irritable bowel syndrome; MT-BC, board-certified music therapist; MBI, music-based intervention; ML, machine learning; OIME index, Objective Integrated Multimodal Electrophysiological (OIME) index; PSS-10, Perceived Stress Scale-10; QST, quantitative sensory testing; 16S rRNA seq, 16S ribosomal RNA gene sequencing.

### Study Design and Setting

This study is a single-arm, prospective pilot mechanistic trial of a four-week MBI in adults with IBS. The study consists of a baseline lab visit, a four-week home-based MBI period, a post-intervention lab visit at Week 5, and follow-up online surveys at Weeks 8 and 12. Lab procedures will be conducted at a biobehavioral research laboratory at the Yale School of Nursing Biobehavioral Research Laboratory in Orange, Connecticut. The study protocol was approved by the Yale University Institutional Review Board (IRB # 2000039033) and registered at ClinicalTrials.gov (NCT06706778). All participants will provide electronic informed consent before enrollment and study procedures begin.

### Participants

Participants will be eligible if they: (1) are 18-50 years of age; (2) have a healthcare provider-confirmed diagnosis of IBS; (3) report an average abdominal pain intensity of ≥3 on a 0-10 numeric rating scale during the past three months; (4) are able to read and communicate in English; (5) are willing to participate in the four-week intervention and attend two lab visits; and (6) have internet access to complete required study procedures.

Individuals will be excluded if they: (1) have a severe psychiatric disorder requiring inpatient treatment within the past six months; (2) regularly use opioids or illicit substances; (3) have used antibiotics or probiotics within two weeks before enrollment; (4) have celiac disease, inflammatory bowel disease, or a history of major gastrointestinal surgery; or (5) are concurrently participating in another IBS-related intervention study.

### Sample Size

A target sample of 36 participants was selected to account for an anticipated attrition rate of approximately 15-20%, yielding an expected sample of approximately 30 participants completing the Week 5 post-intervention lab visit. The sample size was determined primarily by the pilot nature of the study (Hertzog, 2008) and informed by the research team’s prior experience conducting mechanistic assessments in individuals with IBS (Cong, Ramesh, et al., 2018). The anticipated sample is intended to provide preliminary estimates of feasibility and clinical and mechanistic responses to inform the design, measurement strategies, and sample size planning of a future adequately powered trial. Supplementary Appendix S1 provides additional details on sample size considerations and the statistical analysis plan.

### Music-Based Intervention

#### Intervention Content and Structure

Based on the NIH Music-Based Intervention Toolkit (Edwards et al., 2023) and reporting guidelines (Robb et al., 2025), the intervention music and delivery procedures were selected and refined in collaboration with a music research expert and a board-certified music therapist. The intervention uses a single, standardized recording performed on a 25-string monochord (Musicmakers, Stillwater, MN), tuned entirely to C in the Low C configuration: 23 bronze-wound strings (.036” gauge) tuned to C3, and 2 bronze-wound strings (.060” gauge) tuned one octave lower to C2. The instrument is played using a continuous alternating-hand strumming technique across the full string set, producing a sustained, seamless drone rather than discrete struck notes. The same recording is used across all lab and home sessions for all participants. Acoustic analysis of the recording confirmed a sustained low-frequency fundamental of approximately 131 Hz (C3, present in over 93% of pitched frames), consistent with the instrument’s tuning, along with rich harmonic overtone content and stable spectral characteristics with minimal pitch drift throughout. These acoustic characteristics provide a stable, minimally varying auditory stimulus intended to support relaxation and pain modulation. Prior studies of music listening and monochord-based stimulation have also reported associations with relaxation-related responses and changes in electroencephalogram (EEG) theta activity (Ara & Marco-Pallares, 2021; Martin-Saavedra et al., 2018; Sandler et al., 2017; Sandler et al., 2016); accordingly, theta activity is examined in the present study as a candidate neurophysiological response rather than an established property of the intervention.

Each MBI session follows a standardized 20-minute structure consisting of a 5-minute pre-MBI relaxation period, a 10-minute music-listening period, and a 5-minute post-MBI recovery period with quiet rest (Figure 2). Participants will be instructed to listen using a comfortable volume through headphones in a quiet environment, while wearing the designated wearable devices during each session. Multimodal physiological signals and real-time pain ratings will be collected continuously across the entire 20-minute period.

**Figure 2.**
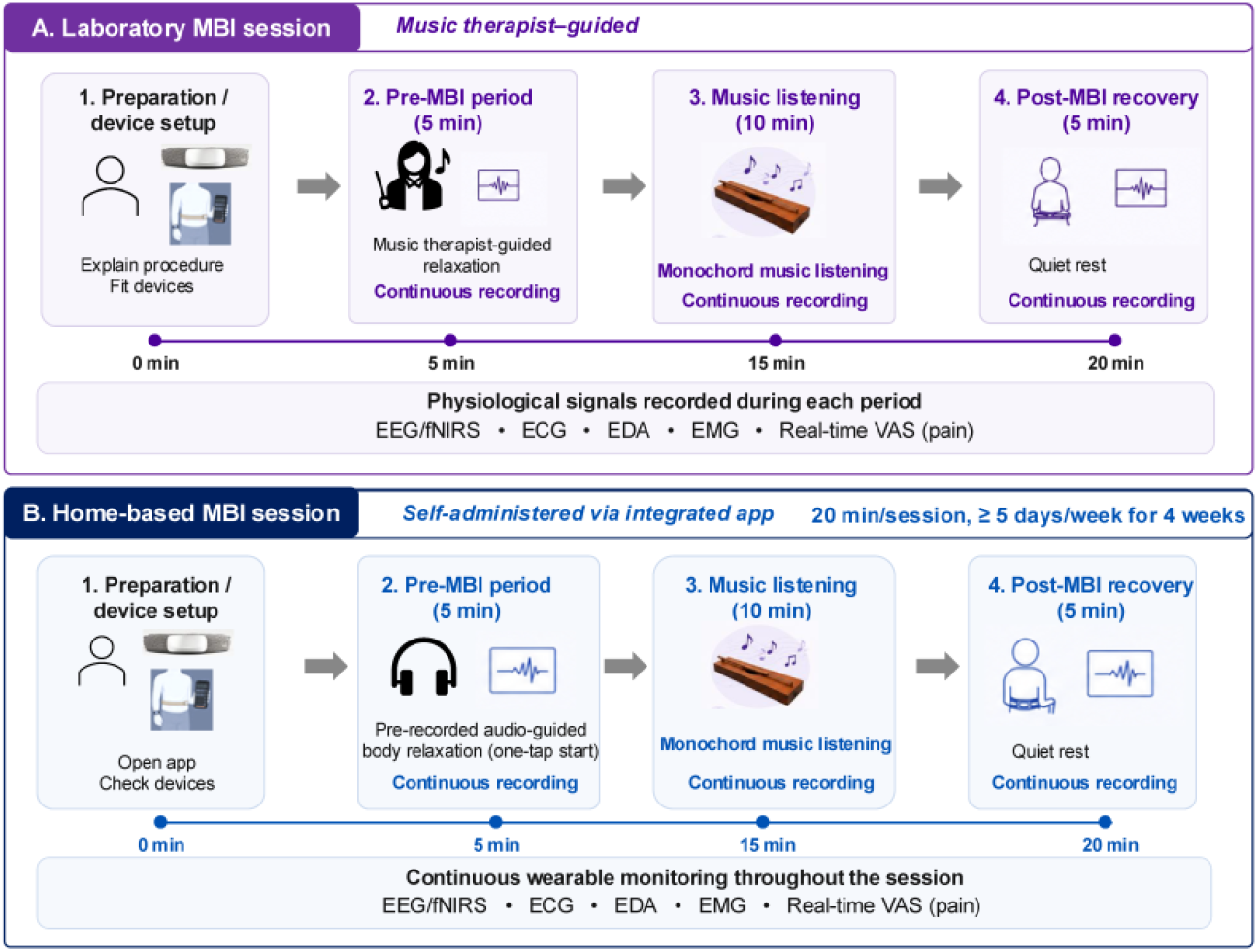
Standardized lab and home-based music-based intervention procedures. Lab sessions include a 5-min therapist-guided relaxation period, 10-min monochord music-listening period, and 5-min recovery period, with continuous multimodal physiological monitoring. Home sessions follow the same standardized intervention sequence, with the relaxation component delivered through a prerecorded audio guide, using the study app and wearable devices.

#### Laboratory Delivery and Fidelity

Consistent with recommendations from the NIH Behavior Change Consortium (Bellg et al., 2004) and their application to music-based interventions (MacLean et al., 2022; Robb et al., 2011), standardized procedures will be used to support treatment fidelity across laboratory and home delivery. During lab sessions, a master’s-prepared, board-certified music therapist (MT-BC) with specialized training and clinical experience in relaxation and MBI delivery will provide participants with intervention orientation and guide the pre-MBI relaxation period. The MT-BC and research staff will complete protocol-specific training before study implementation. The same standardized recording, 20-minute session structure, listening instructions, and wearable-monitoring procedures will be used for all participants at both the baseline and Week 5 lab visits to support consistent intervention delivery.

#### Home Delivery and Fidelity

For home practice, the same standardized session structure is maintained, with the relaxation component delivered through a prerecorded audio guide. After the baseline lab session, participants will be instructed to complete at least one 20-minute MBI session per day, at least five days per week, for four weeks. Participants may complete additional sessions, particularly during episodes of increased abdominal pain or gastrointestinal discomfort, to support pain and symptom self-management.

The study-specific application will standardize home intervention delivery and automatically record session initiation, duration, and completion, while the synchronized wearable system will capture physiological data acquisition. These data will be supplemented by participant-reported session frequency and timing in REDCap diaries to characterize adherence and fidelity of home-based intervention enactment. A home MBI session will be considered complete when participants complete the full app-guided 20-minute intervention sequence.

Home MBI adherence will be quantified based on completion of the minimum 20 prescribed sessions over four weeks and the number of weeks in which participants complete at least five sessions; additional sessions will be recorded separately. Study personnel will conduct weekly follow-up contacts to address technical issues, reinforce adherence, and answer participant questions. Because this is a single-arm pilot without a comparison condition, treatment differentiation and contamination between conditions are not applicable (Reschke-Hernandez & Tranel, 2024).

### Recruitment

Participants will be recruited through multiple complementary strategies, including collaborating gastroenterology practices within Yale New Haven Health, community-based advertisements, social media and online IBS communities, and institutionally approved electronic recruitment approaches. Potentially eligible individuals may also be identified through IRB-approved electronic health record-based procedures, including MyChart outreach and limited review of electronic health records by authorized study personnel. Individuals who express interest will complete an initial self-eligibility screening, and trained study personnel will verify their responses. Those who meet the eligibility criteria will receive a detailed explanation of the study procedures, rights and risks, and expectations, and will provide electronic informed consent (eConsent) before enrollment and initiation of any study-specific procedures.

### Study Procedures

Figure 3 summarizes the overall participant timeline and schedule of study assessments. Following enrollment, participants will complete a baseline survey and lab visit before initiating the four-week home-based intervention. At this visit, participants will receive standardized training in the MBI and the use of study wearable devices and complete their first supervised 20-minute MBI session. Quantitative Sensory Testing (QST) assessments will be conducted before and after the MBI session. Participants will also receive a stool collection kit and standardized instructions for home sample collection.

**Figure 3.**
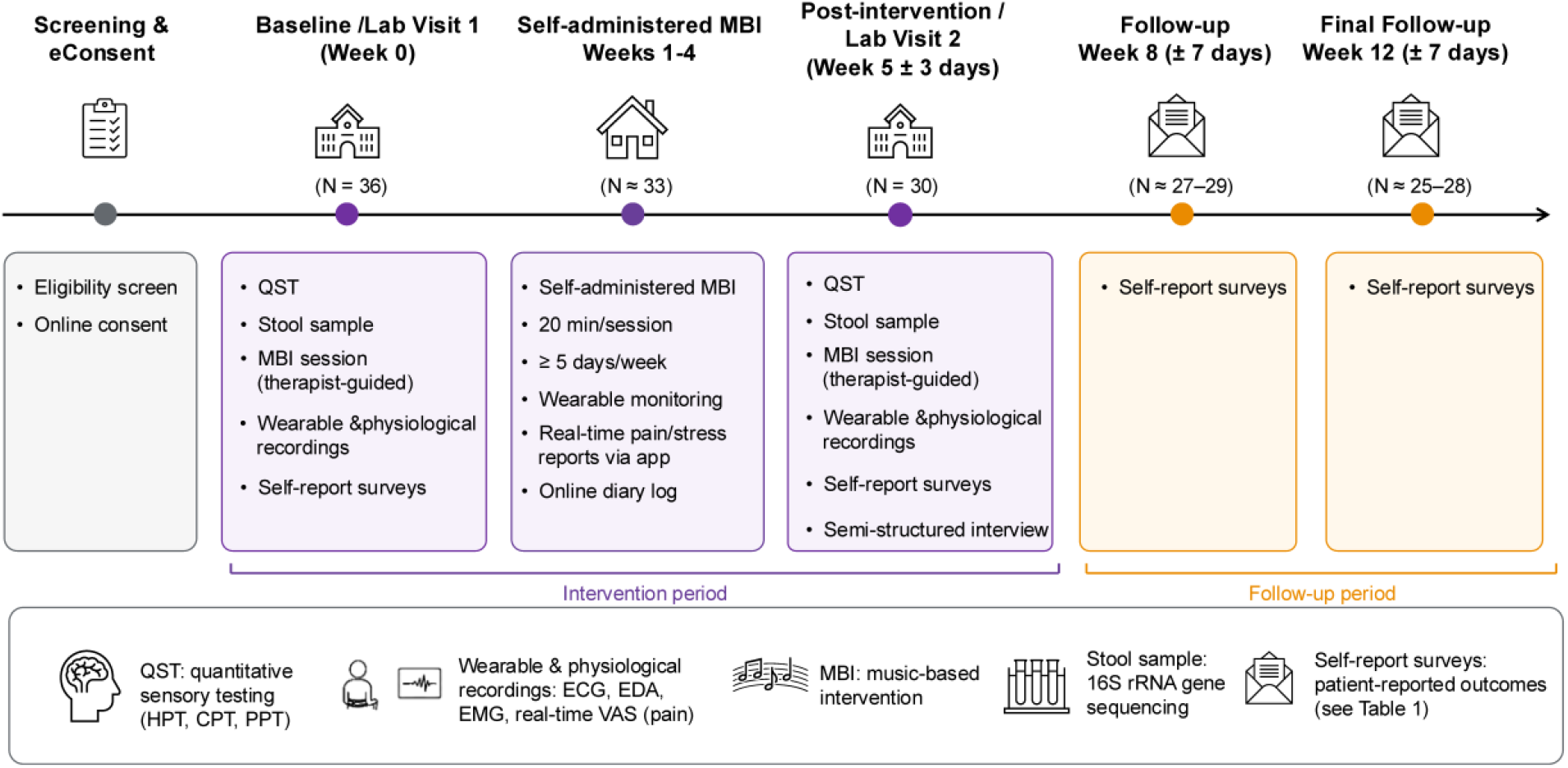
Study flow and assessment schedule. Participants will complete screening and enrollment, a baseline lab visit, four weeks of home-based MBI practice, a post-intervention laboratory visit, and follow-up assessments at Weeks 8 and 12. Abbreviations: MBI, music-based intervention; QST, quantitative sensory testing; EEG, electroencephalography; fNIRS, functional near-infrared spectroscopy; ECG, electrocardiography; EDA, electrodermal activity; EMG, electromyography; VAS, visual analog scale.

During the four-week home-based intervention period, participants will use the study-specific app to initiate each standardized 20-minute MBI session and synchronized wearable recording through a single-step interface to support consistent implementation in the home environment.

At Week 5, participants will return for a post-intervention lab visit following the same standardized MBI and multimodal assessment protocol used at baseline. Participants will also complete a 30-minute semi-structured qualitative interview on intervention acceptability and their experience using the wearable technologies. Follow-up surveys will be conducted remotely at Weeks 8 and 12.

To maximize participant retention, reminder notifications will be sent before scheduled study visits, and participants will receive study compensation after each lab visit.

### Measures

Overall measures include sociodemographic, clinical, and music background data; patient-reported pain-related outcomes; physiological signals; somatic pain sensitivity assessments; gut microbiome profiling; and feasibility indicators. Table 1 summarizes the study assessments and data collection schedule.

**Table 1.**
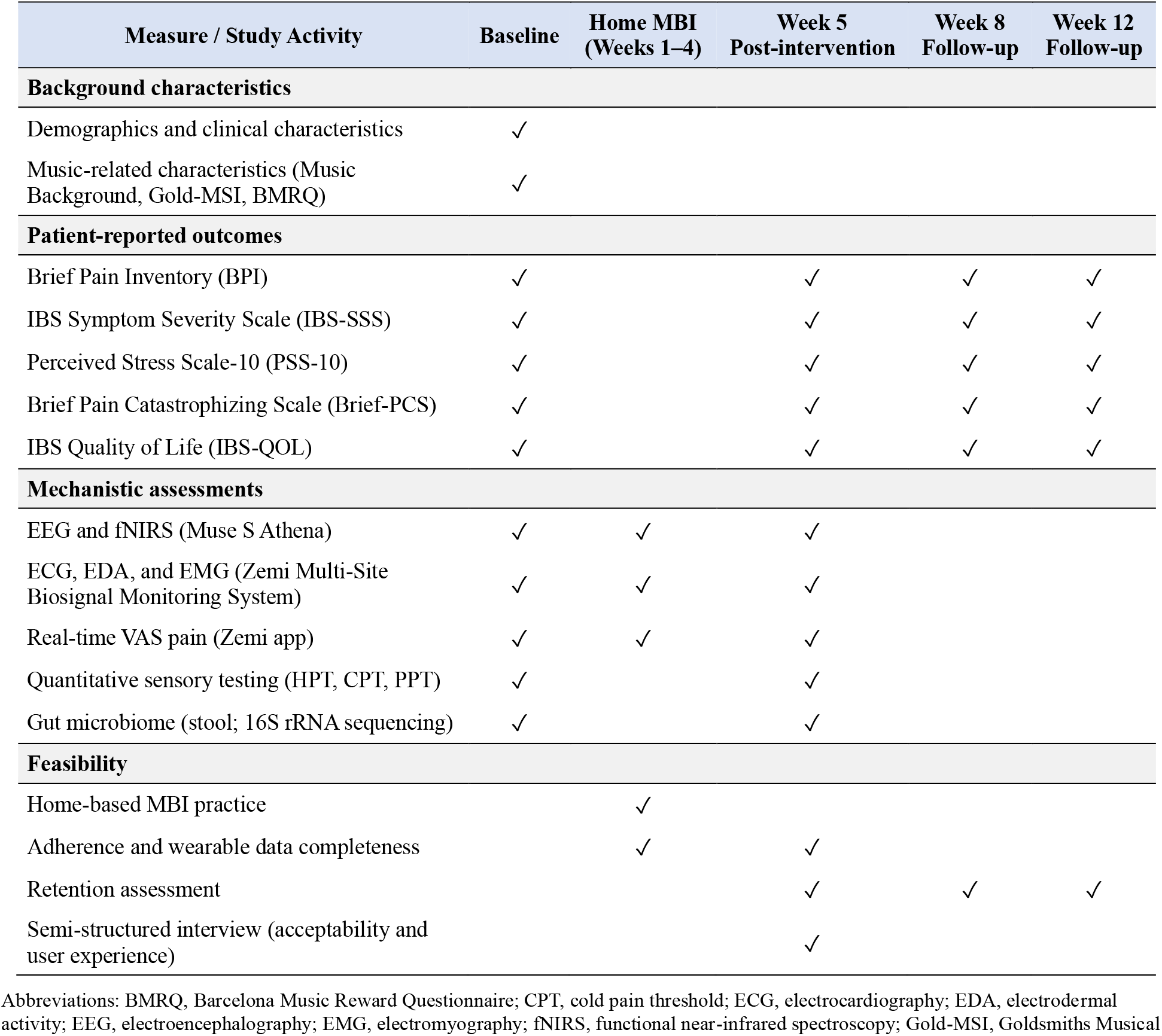
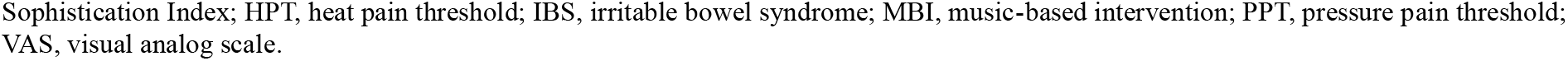
Schedule of study assessments and data collection.

#### Sociodemographic and Clinical Characteristics

A brief baseline questionnaire will collect sociodemographic and clinical characteristics, including age, biological sex, gender, race, ethnicity, education level, employment status, marital status, IBS diagnosis and subtype, pain history, pharmacological treatment, and the number of healthcare visits for IBS-related pain during the previous six weeks.

#### Music-Related Characteristics

**Musical sophistication** will be assessed at baseline using the Goldsmiths Musical Sophistication Index (Gold-MSI) (Mullensiefen et al., 2014), a validated self-report measure of individual differences in musical engagement and abilities in the general population. The Gold-MSI assesses multiple dimensions of musical sophistication, including active engagement, perceptual abilities, musical training, singing abilities, and emotional responses to music, and has demonstrated good reliability and validity (Mullensiefen et al., 2014). Additional baseline questions will assess music preferences, listening habits, and prior use of music for symptom management.

**Music reward sensitivity** will be assessed using the Barcelona Music Reward Questionnaire (BMRQ) (Lippolis et al., 2025; Mas-Herrero et al., 2013), a validated self-report measure of individual differences in the rewarding experiences associated with music. The BMRQ assesses five dimensions of music reward: musical seeking, emotion evocation, mood regulation, social reward, and sensory-motor responses. Baseline music reward sensitivity will be explored as an individual characteristic potentially associated with engagement in the MBI and subsequent clinical and physiological responses.

#### Patient-Reported Pain, Stress, and IBS-Related Outcomes

**Pain severity and pain interference** will be assessed using the Brief Pain Inventory (BPI) (Keller et al., 2004), a validated measure widely used in chronic pain research. The BPI includes four items assessing pain severity and seven items assessing pain interference with daily functioning, each rated on a numeric rating scale from 0 to 10, with higher scores indicating greater pain severity or interference. The BPI has demonstrated good internal consistency, with reported Cronbach’s α values ranging from 0.77 to 0.91(Keller et al., 2004).

**IBS symptom severity** will be assessed using the Irritable Bowel Syndrome Severity Scoring System (IBS-SSS) (Francis et al., 1997). The IBS-SSS assesses abdominal pain severity and frequency, abdominal distension, dissatisfaction with bowel habits, and interference with daily life, generating a total score ranging from 0 to 500, with higher scores indicating greater symptom severity. The instrument has demonstrated good psychometric properties and responsiveness to changes in IBS symptoms (Francis et al., 1997).

**Pain catastrophizing** will be assessed using the four-item Brief Pain Catastrophizing Scale (Brief-PCS) (Walton et al., 2020), a validated abbreviated measure of maladaptive cognitive and emotional responses to pain. Items are rated on a 5-point scale from 0 (not at all) to 4 (all the time), yielding a total score ranging from 0 to 16, with higher scores indicating greater pain catastrophizing. The abbreviated scale efficiently assesses pain-related catastrophic thinking while reducing participant burden.

**Perceived stress** will be assessed using the 10-item Perceived Stress Scale-10 (PSS-10) (Cohen et al., 1983), which measures the extent to which situations in one’s life are perceived as unpredictable, uncontrollable, and overwhelming. Items are rated on a 5-point scale from 0 (never) to 4 (very often), with higher scores indicating greater perceived stress. The PSS-10 has demonstrated good internal consistency and test-retest reliability (Cohen et al., 1983).

**IBS-related quality of life** will be assessed using the 34-item Irritable Bowel Syndrome Quality of Life questionnaire (IBS-QOL) (Hahn et al., 1997). Items are rated on a 5-point scale and transformed to a 0-100 scale, with higher scores indicating better IBS-specific quality of life. The IBS-QOL has demonstrated excellent internal consistency (Cronbach’s α = 0.96), construct validity, and responsiveness to changes in IBS symptoms (Andrae et al., 2013; Drossman et al., 2000).

#### Physiological Measures

**Physiological activity** will be continuously recorded using wearable devices during lab MBI sessions and home-based practice. Central neural activity will be assessed using the Muse S Athena headband (*Muse™ S Athena Headband Manual*), which simultaneously records electroencephalography (EEG) and functional near-infrared spectroscopy (fNIRS). EEG will characterize cortical oscillatory activity associated with music processing and pain modulation, with particular exploratory attention to theta-band activity as a candidate neural response to the MBI. fNIRS will quantify cortical hemodynamic responses related to cognitive, emotional, and pain processing (Chen & Sun, 2017; Hu et al., 2021; Uchitel et al., 2021). Peripheral physiological activity will be recorded using two devices developed by Zemi Labs Inc.: a wearable abdominal monitoring belt that integrates electrocardiography (ECG), electrodermal activity (EDA), and electromyography (EMG), and a wrist-based monitor for palmar EDA. Together, the belt and wrist device constitute a *Zemi Multi-Site Biosignal Monitoring System*. ECG-derived heart rate and heart rate variability (HRV) will characterize cardiac autonomic regulation (Malik, 1996); EDA will index sympathetic arousal (Boucsein et al., 2012); and EMG will capture abdominal muscle activity. Together, these synchronized recordings will enable multimodal characterization of physiological responses before, during, and following MBI exposure.

#### Quantitative Pain Sensitivity

Pain sensitivity will be assessed using a standardized QST protocol adapted from the German Research Network on Neuropathic Pain (Rolke et al., 2006; Weaver et al., 2021). QST will be conducted immediately before and after the MBI session at both lab visits, enabling assessment of acute within-session changes in pain sensitivity as well as changes from baseline to post-intervention following the 4-week MBI. Heat pain threshold (HPT), cold pain threshold (CPT), and pressure pain threshold (PPT) will be assessed at two anatomical sites: the participant-identified abdominal pain site and the non-dominant forearm as a remote comparison site. Testing at the abdominal site will characterize pain sensitivity in the symptomatic region, whereas the forearm will provide an index of pain sensitivity remote from the primary pain site (Verne et al., 2003). Together, within-session pre-post comparisons, between-visit changes, and differences between anatomical sites will be used to characterize acute and longer-term patterns of pain modulation associated with the MBI.

#### Gut Microbiome

**Gut microbiome composition** will be assessed using stool samples collected at baseline and post-intervention with the researcher-provided OMNIgene●GUT self-collection kit (DNA Genotek, Inc.), which stabilizes samples at room temperature prior to processing (Chen et al., 2020). Participants will receive standardized instructions for self-collection during lab visits and return samples using prepaid mailing materials (Szopinska et al., 2018). Samples will be stored at −80°C until processing. Microbial DNA will be extracted from stool samples, and the V4 region of the 16S rRNA gene will be amplified and sequenced using paired-end sequencing on an Illumina MiSeq platform at the University of Connecticut Microbial Analysis, Resources, and Services facility. Sequencing reads will undergo standardized quality filtering and preprocessing using the established MiSeq analysis pipeline before taxonomic profiling and diversity analyses. The research team has previously implemented this stool collection, sequencing, and analytic workflow in adults with IBS (Cong, Ramesh, et al., 2018; Wu et al., 2025).

#### Feasibility of Home-Based MBI and Wearable Monitoring

Feasibility will be evaluated using both quantitative and qualitative indicators, including recruitment, retention, home MBI adherence, wearable-device adherence, physiological data completeness, and participant acceptability. Home MBI adherence will be summarized using the prespecified session-completion and adherence criteria described above. Wearable-device adherence will be assessed based on device use during prescribed home sessions, supplemented by participant-reported frequency, timing, and duration of device use recorded in structured REDCap diaries. Physiological data completeness will be evaluated based on successful device synchronization and the proportion of usable versus missing data across recording modalities.

Retention will be assessed based on participant completion of scheduled study assessments. Post-intervention semi-structured interviews will be conducted to assess participant experiences, barriers to home implementation and wearable use, and overall acceptability.

### Data Analysis

All statistical analyses will be performed in R. All enrolled participants will be included in study-flow and feasibility summaries, and outcome-specific analyses will use measurements that meet prespecified scoring and quality criteria. Descriptive statistics will be used to summarize participant characteristics, feasibility, and outcome measures. Continuous variables will be summarized using means and standard deviations or medians and interquartile ranges, as appropriate, and categorical variables will be summarized using frequencies and percentages with explicit denominators. Missing data patterns will be described and examined. Likelihood-based mixed-effects models will use all available valid repeated observations. Multiple imputation will be conducted if warranted by the extent and pattern of missingness. When appropriate, multivariable models will adjust for prespecified participant characteristics, including age, sex, baseline pain severity, and other clinically relevant variables to examine their potential influence on observed outcome trajectories. Supplementary Appendix S1 provides detailed statistical assumptions, model specifications, and sensitivity analyses.

#### Clinical Outcomes

The primary outcome is BPI pain severity, the primary time point is Week 5, and the primary estimand is the mean within-participant change from baseline to Week 5. BPI pain severity measured at baseline and Weeks 5, 8, and 12 will be analyzed using a linear mixed-effects model with categorical assessment time as a fixed effect and a participant-specific random intercept. The primary Week-5-minus-baseline contrast will be reported in BPI points with a 95% confidence interval and two-sided P value. Baseline-to-Week-8 and baseline-to-Week-12 contrasts will assess maintenance. BPI pain interference, IBS symptom severity, quality of life, perceived stress, and pain catastrophizing will be analyzed using analogous outcome-specific models and treated as secondary outcomes. The single primary comparison will use a two-sided alpha level of .05.

#### Mechanistic Outcomes

Mechanistic analyses will characterize changes in pain sensitivity, physiological activity, and gut microbiome profiles associated with the MBI. For QST outcomes, changes in HPT, CPT, and PPT will be examined across lab visits (baseline and post-intervention), within-session periods (pre- and post-MBI), with repeated measurements accounted for using mixed-effects models. Physiological recordings, including EEG, fNIRS, ECG, EDA, and EMG, will be processed to derive modality-specific time- and frequency-domain features and to characterize physiological responses across pre-music, music-listening, and post-music recovery periods. A previously developed machine learning/deep learning-based Objective Integrated Multimodal Electrophysiological (OIME) index will be applied to the ECG, EDA, and EMG data collected in this study to generate an objective estimate of IBS pain intensity. OIME-derived pain estimates will be compared with concurrent real-time VAS pain ratings to explore the index’s performance and potential utility for objective pain monitoring in adults with IBS. Functional regression and other longitudinal modeling approaches will be used, as appropriate, to explore associations between continuous physiological responses and clinical outcomes.

For gut microbiome outcomes, sequencing data will undergo standardized quality-control and preprocessing procedures before downstream analysis. Alpha diversity will be characterized using the Shannon index, inverse Simpson index, and observed richness, and beta diversity will be characterized using Bray-Curtis dissimilarity. Within-participant changes in alpha diversity from baseline to post-intervention will be evaluated using paired parametric or nonparametric tests, as appropriate, while changes in community composition will be assessed using PERMANOVA accounting for the paired study design. Exploratory taxonomic analyses will examine longitudinal changes in microbial relative abundance, with false discovery rate correction applied to multiple comparisons. Given the pilot sample size, microbiome analyses will be considered exploratory and hypothesis-generating.

#### Exploratory Mechanism-Outcome Associations

Exploratory analyses will examine whether changes in candidate brain-gut mechanisms, including physiological, sensory, stress-related, and gut microbiome measures, are associated with changes in pain outcomes. Given the pilot sample size, these analyses are intended to characterize patterns of association and generate hypotheses for future mechanistic testing rather than formally establish mediation.

## Discussion

### Study Progress

At the time of manuscript preparation, the study was in the final preparatory phase before participant recruitment. Study protocols, standard operating procedures, case report forms, and data management systems have been finalized, and research staff, including the music therapist and lab personnel, have completed protocol-specific training to support standardized MBI delivery and data collection. Final preparation, calibration, and validation of the wearable abdominal monitoring system are underway. Participant recruitment will begin once these final device-related procedures are completed.

### Lessons Learned

A key lesson from study preparation is the need to integrate multimodal wearable technologies into a participant-friendly workflow suitable for both lab and home settings. The study requires synchronized physiological recordings from the Zemi Multi-Site Biosignal Monitoring System (belt and wrist devices) and a commercially available Muse S Athena headband while minimizing the technical burden placed on participants. To address this challenge, Zemi Labs worked with the music therapist and study investigators to streamline intervention delivery and multimodal physiological recording into a single-step workflow within the Zemi mobile app. With one participant-initiated action, the application launches the standardized MBI sequence and synchronized wearable recording, reducing the need for participants to operate multiple systems independently. This integrated approach is intended to reduce operational complexity and recording errors while supporting intervention adherence and physiological data completeness during home-based practice. This experience highlights the importance of considering participant-facing workflow, rather than device capability alone, when translating multimodal physiological monitoring from controlled laboratory environments to repeated home use.

A second methodological lesson concerns the challenge of capturing physiological responses during clinically meaningful IBS pain. Because IBS pain is episodic and varies both within and between individuals, participants may present for lab assessments with minimal or no active abdominal pain, limiting the ability to characterize pain-related physiological responses and their modulation during the MBI. To increase the likelihood of capturing symptomatic periods, lab visits will be scheduled, whenever feasible, according to each participant’s typical temporal pattern of IBS pain. If active pain is minimal at the scheduled visit, a standardized cold-water ingestion challenge may be used to elicit gastrointestinal symptoms (Zuo et al., 2006), consistent with the study protocol. A participant-identified dietary trigger known from prior experience to provoke IBS symptoms may also be considered, contingent on IRB approval and participant consent (Shepherd et al., 2008). Pain intensity will be reassessed following symptom elicitation before proceeding with the MBI protocol. In parallel, repeated home-based physiological monitoring provides an opportunity to capture MBI-related responses during naturally occurring episodes of increased abdominal pain or gastrointestinal discomfort. Participants may also complete additional intervention sessions during symptomatic periods, with concurrent wearable monitoring and real-time pain assessment enabling characterization of physiological responses under more ecologically relevant conditions. Together, these strategies illustrate the value of combining standardized lab assessments with repeated real-world monitoring when studying mechanisms underlying episodic and fluctuating symptoms such as IBS pain.

A third implementation consideration is participant recruitment and retention. The multimodal and longitudinal nature of the protocol may increase participant burden and limit the pool of individuals willing and able to complete the study. Based on recruitment experience from related studies conducted by the research team, sustained and flexible recruitment efforts may therefore be necessary to achieve the target sample. To address this anticipated challenge, the study will use a multimodal recruitment approach spanning clinical, institutional, community, and digital sources, adjusting recruitment intensity based on enrollment patterns and seasonal availability of the target population. The study will support participant retention through streamlined procedures, regular communication, reminders, and technical assistance throughout the intervention. Recruitment, retention, and adherence metrics collected during this pilot will help determine whether these strategies are sufficient and identify refinements needed for a future larger-scale trial.

### Limitations

Several limitations of this pilot study should be acknowledged. The single-arm design and modest sample size preclude definitive conclusions regarding intervention efficacy or causal effects. Given the breadth of clinical and mechanistic assessments, analyses are primarily exploratory and hypothesis-generating, and associations between mechanistic responses and changes in pain cannot establish causal or mediating pathways. In addition, variability in adherence to home-based intervention and wearable-monitoring procedures may contribute to heterogeneity in intervention exposure and data completeness; these factors will be explicitly characterized as part of the feasibility evaluation. These limitations are consistent with the pilot nature of the study, which is intended to identify candidate mechanistic signals and inform the design, outcome selection, and analytic priorities of a future adequately powered randomized controlled trial.

### Potential Impact

This pilot study provides a mechanistically informed approach to evaluating MBI for IBS-related pain. By integrating patient-reported outcomes with multimodal physiological, sensory, and gut microbiome measures, the study will enable characterization of clinical and candidate mechanistic responses across multiple domains relevant to IBS pain. The integration of laboratory assessments with repeated home-based wearable monitoring may further provide insight into the feasibility of capturing physiological responses across both controlled and everyday settings. Findings will inform the refinement of mechanistic hypotheses, outcome and measurement strategies, and implementation procedures for future adequately powered trials, with the longer-term goal of advancing mechanism-informed, nonpharmacological approaches to IBS symptom management.

## Acknowledgements

The authors acknowledge the University of Connecticut Microbial Analysis, Resources, and Services (MARS) facility for its support with 16S rRNA gene sequencing and microbiome analyses. The authors also thank the Yale School of Nursing Biobehavioral Laboratory for its support in study preparation, protocol development, and implementation.

## Ethics Statement

The study protocol was approved by the Yale University Institutional Review Board (IRB #2000039033). All participants will provide electronic informed consent before enrollment and initiation of any study-specific procedures. The study is registered at ClinicalTrials.gov (NCT06706778).

## Data Availability Statement

Data sharing is not applicable to this article because no datasets were generated or analyzed for the preparation of this protocol manuscript. Data generated from the study will be managed and shared in accordance with the approved study protocol, institutional requirements, and applicable data-sharing policies.

