## Supplementary Appendix S1 for "Clinical and mechanistic responses to a music-based intervention for pain in adults with irritable bowel syndrome: Protocol for a single-arm pilot study" (from another version of this preprint: version 1, 2026-09-17)

**Detailed Data Analysis Plan for the Pilot Study**

**General Statistical Procedures**

Self-reported data will be exported from the REDCap database for analysis. All electrophysiological signal readings will be downloaded into the online log weekly. Data-quality checks will address completeness, plausible ranges, internal consistency, duplicate records, temporal alignment, and modality-specific signal quality. R (currently version 4.6.1, or the most updated version available at the time of analysis) will be used for data analysis. All enrolled participants will be included in study-flow and feasibility summaries. Outcome-specific analyses will use measurements meeting prespecified scoring and quality criteria, with the number of participants and observations contributing at each assessment reported.

Continuous variables will be summarized using means and standard deviations or medians and interquartile ranges, as appropriate, and categorical variables using frequencies and percentages with explicit denominators. The amount and pattern of missing data will be described by outcome and assessment. Likelihood-based mixed-effects models will use all available valid repeated observations under a missing-at-random assumption conditional on variables included in the model. Multiple imputation will be conducted if warranted by the extent and pattern of missingness. The single primary comparison is the Week-5-minus-baseline change in Brief Pain Inventory (BPI) pain severity and will be evaluated at a two-sided alpha level of .05 without multiplicity adjustment. Other clinical contrasts and mechanistic analyses are secondary or exploratory; estimates and 95% confidence intervals will be emphasized. Model evaluation will address convergence and singularity, residual distributions and variance assumptions, influential observations, and adequacy of the repeated-measure covariance structure; alternative covariance structures will be considered only as sensitivity analyses when estimable.

**Aim 1: Explore changes in IBS pain and candidate mechanisms observed during the MBI study**

**1.1 Clinical and mechanistic outcome analyses**

The primary outcome is BPI pain severity, the primary time point is Week 5, and the primary estimand is the mean within-participant change from baseline to Week 5. BPI pain severity measured at baseline and Weeks 5, 8, and 12 will be analyzed using a linear mixed-effects model with assessment time represented categorically as a fixed effect, baseline as the reference assessment, and a participant-specific random intercept. Models will be estimated using restricted maximum likelihood, with Satterthwaite degrees of freedom used for confidence intervals and testing. The primary Week-5-minus-baseline contrast will be reported in BPI points with a 95% confidence interval and two-sided P value. Baseline-to-Week-8 and baseline-to-Week-12 contrasts will characterize maintenance. The primary model will be unadjusted. Parsimonious covariate-adjusted analyses may be conducted as sensitivity analyses using a small prespecified set of baseline characteristics, such as age and biological sex, if model stability permits. Baseline BPI pain severity will not be entered as a separate covariate because it is included as part of the repeated outcome trajectory. BPI pain interference, IBS symptom severity, IBS-related quality of life, perceived stress, and pain catastrophizing will be analyzed using analogous outcome-specific models and treated as secondary outcomes.

Heat, cold, and pressure pain thresholds will be analyzed in separate mixed-effects models. Fixed effects will include laboratory visit (baseline and Week 5), within-session period (pre- and post-MBI), anatomical site (abdominal pain site and nondominant forearm), and the visit-by-period interaction; participant will be included as a random intercept. The visit-by-period contrast will estimate whether the acute pre-to-post response differs between baseline and Week 5. Site-specific contrasts and the visit-by-period-by-site interaction will be exploratory and fitted only if model stability permits. Outcomes affected by marked skewness, influential values, or instrument limits will be evaluated using an appropriate transformation, robust procedure, or paired nonparametric sensitivity analysis.

Stool samples collected at baseline and Week 5 will be analyzed as paired observations. Within-participant changes in alpha diversity, including Shannon diversity and Chao1 richness when estimable, will be summarized using paired t tests or Wilcoxon signed-rank tests, as appropriate. Community composition will be evaluated using prespecified distance measures, including Bray–Curtis and UniFrac distances when available, and PERMANOVA with permutations constrained within participant; multivariate dispersion will be assessed separately. Exploratory taxonomic differential-abundance analyses will be conducted using ANCOM-BC2, whereas microbial functional profiles will be computationally predicted using Tax4Fun2. Analyses will account for the paired baseline and Week 5 design, with feature filtering, normalization or transformation, covariate adjustment, and false-discovery-rate control specified separately for the taxonomic and predicted functional feature families. Exploratory analyses may relate changes in physiological, sensory, stress-related, or microbiome measures to changes in BPI pain severity using parsimonious change-score or longitudinal models.

**1.2 Functional regression of physiological and neurophysiological trajectories**

Functional regression, implemented using the fda package in R or an equivalent validated implementation, will be used to examine associations between changes in clinical outcomes, such as BPI pain severity and PSS-10, and continuous physiological and neurophysiological trajectories recorded during the 20-minute MBI session. These trajectories may include heart rate variability derived from ECG, as well as EDA, EMG, EEG, and fNIRS measures. Each participant's time-series data will be represented as functional predictors after prespecified signal-quality assessment, smoothing, and feature construction. Functional coefficient estimates will characterize time regions associated with the clinical outcome. Given the pilot sample size, model complexity and covariate adjustment will be limited, and these analyses will be considered exploratory.

**Aim 2: Evaluate the feasibility of integrating multimodal wearable physiological monitoring into repeated home-based MBI sessions.**

Recruitment, retention, intervention adherence, wearable use, data completeness, device failure, and acceptability will be summarized. Wearable completeness will be reported by device, participant, session, and recording period. Feasibility proportions will be reported with 95% confidence intervals when informative, and qualitative acceptability findings will be analyzed separately from the quantitative feasibility estimates.

Weekly adherence trajectories during Weeks 1–4 may be evaluated exploratorily using completed and expected session counts in a binomial mixed-effects model with week as a categorical fixed effect and participant as a random intercept. Baseline predictors such as music-related characteristics or demographic factors will be introduced only in parsimonious exploratory models if supported by the number of observations and model stability. If a proportion model is unstable or inconsistent with the observed distribution, adherence will remain descriptive.

**Sample size justification**

The study will enroll 36 participants, with approximately 30 expected to complete the Week-5 assessment after 15%–20% attrition. The target was selected primarily to evaluate feasibility, estimate outcome variability and the precision of within-participant changes, and refine procedures and planning parameters for a future randomized trial. If 30 of 36 enrolled participants complete Week 5, anticipated retention is 83.3%, with a Wilson 95% confidence interval of approximately 68.1%-92.1%.

Sensitivity for the primary BPI estimand was evaluated using the exact noncentral t distribution for a two-sided paired comparison with alpha=.05 and 30 complete baseline-to-Week-5 pairs. Under these assumptions, 80% power corresponds to a minimum detectable standardized paired-change effect of |d|=0.529. Under the provisional planning assumption that the standard deviation of the paired BPI change is 1.39, this corresponds to a minimum detectable mean change of 0.736 BPI, and the anticipated 95% confidence-interval half-width for the mean change is 0.519 BPI. A provisional scenario with a mean change of -0.71 and a paired-change standard deviation of 1.39 yields d=-0.511 and power of approximately 77.2%.

The paired calculation maps directly to the prespecified baseline-to-Week-5 contrast and requires only the paired-change variance. The primary outcome analysis will nevertheless use the categorical-time mixed-effects model so that all available valid observations can contribute under the stated missing-data assumption. No formal sample-size claim is made for functional regression, physiological prediction, high-dimensional microbiome analyses, or exploratory mechanism-outcome associations.

**Application of the OIME Index**

The Objective Integrated Multimodal Electrophysiological (OIME) index is a previously developed multimodal model derived from EDA, ECG, and EMG signals collected using an earlier version of the abdominal monitoring system. In the present pilot study, the existing OIME model will be applied to eligible physiological recordings to generate continuous estimates of pain-related physiological responses. Model development or retraining is not planned using the present pilot sample. Exploratory analyses will examine the correspondence between OIME-derived estimates and contemporaneous participant-reported pain ratings. Given the pilot sample size, these analyses will be considered exploratory and will not be used to establish model validity or generalizability.
